# Assessment of repeatability of a clinical automated pupillometer

**DOI:** 10.1101/2025.05.03.25326928

**Authors:** Jacob Diaz, Jan Skerswetat, Andrew Browne

## Abstract

**Purpose:** To determine the repeatability of a clinical pupillometer in healthy participants with a 5 and 30^−^minute test^−^retest break for both Swinging-Flashlight and Low-High Luminance approach.

**Methods:** 20 healthy participants (mean age: 27 years ±5; 9 females) placed their heads into the device’s headrest and fixated a central spot. A Swinging-Flashlight approach evoked a pupillary light reflex, stimulating alternatingly 8 times each eye by a brief diffuse white light flash followed by a continuous measurement of constriction and dilation of the direct and consensual pupils. For the Low-High Luminance setting, continuous measurements of pupil diameters for a 5 second low luminance display followed by a 5 second high luminance white light. Pupillary light reflex parameters for each test and each eye were calculated by the device. Repeatability was investigated after a 5-minute break time and during a control experiment for 21 participants after a 30-minute break for each approach in counterbalanced order using Bland-Altman analysis.

**Results:** Overall, many parameters for both Swinging-Flashlight and Low-High Luminance approach showed retest biases for all pupil light reflex parameters after a 5-minute test-retest break. These biases were almost completely reduced after a 30-minute break between test and retest for both approaches. The test-retest variabilities as expressed using the Coefficient of Repeatability was reduced after 30-minutes for the majority of the Low-High Luminance results but not for results of the Swinging-Flashlight method.

**Conclusions:** Clinical pupillometry includes the Swinging Flashlight Test (SFL) and Low-High Luminance assays. In SFL, many pupillary dynamic metrics showed test-retest bias at a 5-minute interval, whereas relative afferent pupillary defect (RAPD) measurements remained unbiased at both 5 and 30-minute intervals. Similarly, Low-High Luminance testing showed minimal bias after 30 minutes but exhibited bias at the 5-minute interval. Thus, when evaluating multiple pupillary parameters in scientific or clinical settings, RAPD is less susceptible to bias, while other metrics require longer intervals between testing and retesting.

## 1. Introduction

The pupillary light reflex (PLR) is an important neurological response that provides critical insights into the health of the autonomic nervous system and the functionality of the optic pathway. Phototransduction in the retina triggers a neurochemical signal that travels through the optic nerve, optic chiasm, and optic tract to the pretectal nucleus, which then relays it bilaterally to the Edinger-Westphal nucleus.^1^ This nucleus sends efferent, parasympathetic signals via the oculomotor nerve, ciliary ganglion, and short ciliary nerves to the sphincter pupillae muscle, causing both pupils to constrict. ^2^ Any defect in this process can result in the PLR not functioning as expected. Relative Afferent Pupillary Defect (RAPD) is a clinical sign of asymmetry in the afferent visual pathway in which a pupil does not respond to light to the same degree as the opposite pupil.^2^ It reflects an imbalance in the signal transmission from the retina to the brain, typically indicating unilateral or asymmetrical damage to the optic nerve or severe retinal disease. Instead of both pupils constricting as normal, there may be reduced or even no constriction when light is shined in the eye.^3^

The presence of an RAPD is most commonly associated with conditions affecting the optic nerve such as ischemic optic neuropathy^4^, optic neuritis^5^ and glaucoma^6^, however, it can also be seen age-related macular degeneration^7^ retinal detachment^8^ or traumatic brain injury^9^. Detecting RAPD is critical for diagnosing these pathologies, as it often presents before structural abnormalities that are visible via imaging modalities. Historically, RAPDs have been assessed using the swinging flashlight (SFL) test, a subjective method where the clinician observes the pupillary response as light is alternated between the two eyes in terms of size and velocity of constriction dilation.^1,6,7,8^ Although this method is widely used, its effectiveness relies on the skill and experience of the examiner. Subtle RAPD cases, especially those in early disease stages, can be difficult to detect, leading to variability in diagnosis.^9,13^ Moreover, the kinetics of pupil constriction and dilation cannot be quantified, but only qualitatively assessed this way. Consequently, there has been growing interest in developing automated, examiner-independent approaches to RAPD detection.^3,10^

Pupillography, a technique that measures the PLR, typically use infrared eye trackers to continuously capture pupil diameters and then to fit a pupil light reflex function to estimate pupillary parameters such as the onset latency, constriction amplitude, and constriction and dilation velocities.^14,15^ Therefore, pupillography provides a more comprehensive assessment of the PLR than the clinician-administered subjective approach. Moreover, pupillography has been shown to be able to detect diabetic retinopathy, ^16^ early-stage age-related macular degeneration ^17^ and other neurological conditions, e.g., Alzheimer’s disease^18^ and therefore, it may be a suitable tool for screening and monitoring of ophthalmic and other neurological conditions.

One such commercially available pupillography device is the EyeKinetix®.^19^ This recently upgraded instrument uses software to assess RAPD in a SFL paradigm and pupil size changes during low and high luminance lighting. The latter assay was developed for ophthalmologists that plan contact lens, intra-ocular lens, or other refractive surgical interventions.^19^

The current study’s goal was to investigate the repeatability for both pupillographic methods of this commercial device. Test-retest reliability is essential for any clinical test to ensure that measurements are reproducible and reflect the true clinical state of a patient.^20^ For pupillography specifically, this reliability is crucial both physiologically—by confirming the stability of neural and autonomic pathways—and psychophysically—by minimizing the impact of participant variability and external factors.^21^ Establishing high test-retest reliability in pupillographic assessments ultimately enhances diagnostic accuracy, aids in monitoring clinical progression, and supports evidence-based decision-making in both research and clinical setting. It is a common problem impeding the development of the automated pupilometer.^22^ Previous work quantified the repeatability of the previous version of this device and its SFL test setting.^23^ The researchers investigated test-retest repeatability using three separate tests that used inter-test break durations that varied from 10 to 20-minutes. Also, they deployed an Intra-Class Coefficient (ICC) analysis for each pupil light reflex parameter provided by the device and found that the outcome parameters showed ICCs ranging between 0.65 to 0.90. It is noteworthy that the mean parameter data after each test indicates a systematic reduction after each subsequent trial, suggestive of retest bias.^23^ In the current study, we evaluated test-retest at 5 and 30 minutes to distinguish whether difference in recovery from the photo stress induced by the device during the test may alter results for the 5-minute interval retest group. Furthermore, ICCs have been criticized as inadequate analysis of repeatability due to ICC’s measuring association between variables while a Bland-Atman analysis measures the variation between tests.^17,25^ Therefore, the current study used Bland-Altman analysis^24^ for each parameter as it captures the test-retest variance with limits of agreement, coefficient of repeatability, and mean bias metrics.

## Methods

Participants were provided with detailed written and verbal information regarding the study in advance. Written informed consent was obtained from all participants prior to their involvement in the study. Ethical approval for the study was granted by the University of California, Irvine’s Institutional Review Board (IRB number: 20195254), in accordance with the ethical standards outlined in the Declaration of Helsinki (1975).

### Participants

Eligible participants were screened using a demographic questionnaire that contained questions related to ocular health, past ocular interventions and current medical conditions and prescriptions to rule out any history of ophthalmic pathology or previous ocular surgeries. Interested participants were asked to abstain from vigorous exercise or caffeine one hour before and during test administration intervals. Distance visual acuities for OD, OS, and OU were estimated for all participants using a self-administered orientation judgment psychophysical task.^26^

### Procedure

After consenting to take part in the research, participants performed visual screening and filled out demographic questionnaires. The main experiment then began by asking the participant to take a seat in a chair and to place their head into the device’s headrest. The experimenter instructed each participant to fixate the center of the crossing lines and to maintain it throughout each trial. The participants were also instructed to not move nor talk during the test and the ambient environment was sustained with no talking or background noises. The method order was counterbalanced between participants, with participants randomly being assigned to begin with either the Low-High Luminance method, or the SFL method.

#### Low-High Luminance method

The luminance function testing begins with a period of 5 seconds of dark adaptation where pupillary dilation is evoked, followed by a 5 second period of constant bright white light to stimulate pupillary constriction.

#### Swinging Flashlight method

Pupillary responses for the SFL test were evoked alternatingly 8 times to each eye by a 167 millisecond diffuse white light flash followed by 2.14 second continuous measurement of the PLR, specifically for the direct and consensual eyes pupil diameters.

The protocol described above was repeated with 21 participants with a 30-minute break interval, 15 of whom were the same in each test group.

## Data analysis

Pupillary response parameters for each test and each eye were captured, analyzed and calculated by the device’s software. Linear correlations were performed using Matlab’s *corrplot* function and applied Kendall method to measure the strength and direction of association between two ranked variables while note assuming a linear relationship or require normally distributed data. Repeatability for each methods outcome parameters were investigated using Bland-Altman analysis (Figures 1 and 2).^24^

**Figure 1:**
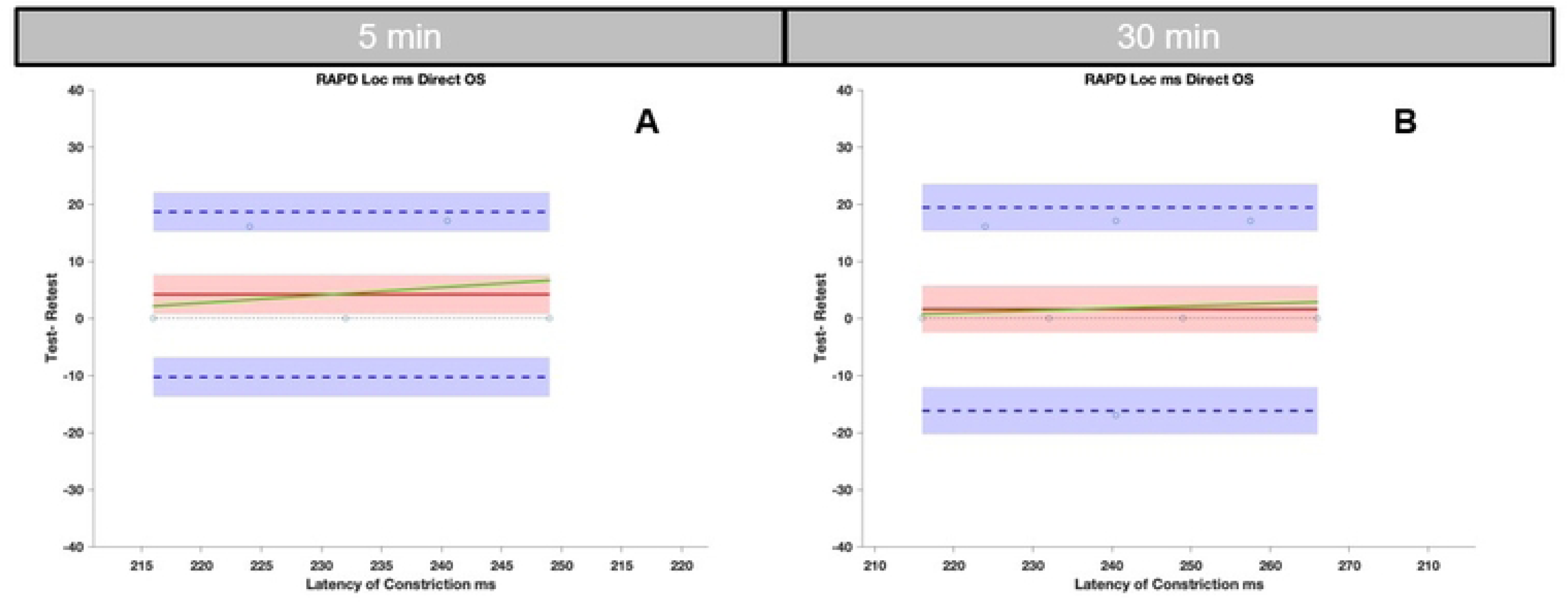
Two example graphs from the SFL setting for the latency of constriction parameter of the direct response in OS. Bland-Altman plots are shown for 5-minute (A) and a 30-minute test-retest interval (B). The black dotted line represents the ideal no bias mark, each circle represented the mean difference between test and retest for each participant. The mean bias and its 95% confidence intervals are represented as the red line with shaded area, respectively. The proportional bias as a function of latency of constriction means is shown as green line. Upper and lower Limits of Agreement are depicted as blue dashed lines with their respective 95% confidence intervals.

**Figure 2:**
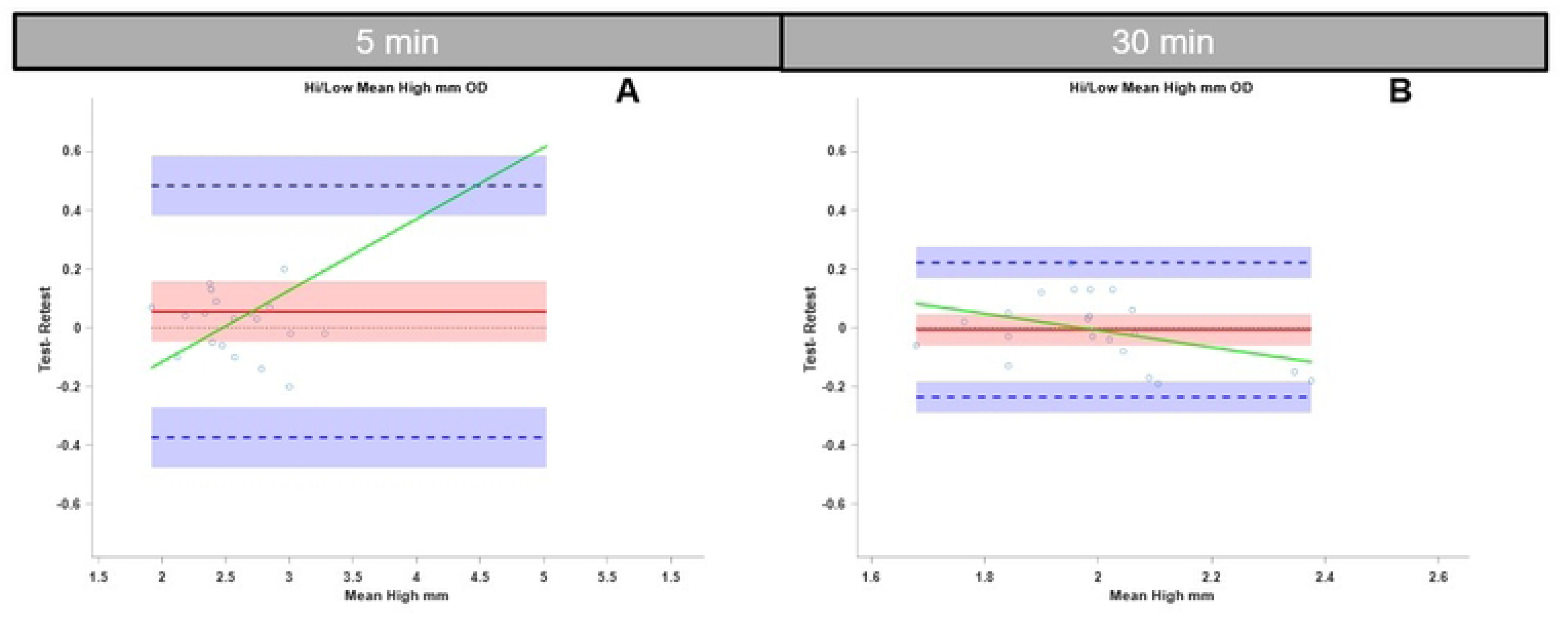
Two example graphs from the Low-High Luminance setting for the High-Low mean parameter in OD. Bland-Altman plots are shown after a 5-minute (A) and 30-minute break (B). The black dotted line represents the ideal no bias mark, each blue circle represented the mean difference between test and retest for each participant. The mean bias and its 95% confidence intervals are represented as the red line with shaded area, respectively. The proportional bias as a function of means is shown as green line. Upper and lower Limits of Agreement are depicted as blue dashed lines with their respective 95% Confidence intervals.

## Results

Means, standard deviations, and Bland-Altman statistics for direct and consensual pupil light reflexes are reported for the 5-minute and 30-minute groups in Appendix Table 1 for SFL method and Appendix Table 2 for the Low-High-Luminance method. Tables 2 and 3 summarize the key observations of biases for SFL test and LHL assays respectively. If the retest tends to give higher or lower values than the first test (instead of being about the same), that shows a *bias toward the retest*. While no test-retest biases were seen for the RAPD assay when evaluated for test-retest intervals of 5 or 30 minutes, many of the dynamic pupillography metrics did show biases for the 5-minute test-retest interval.

**Table 1:**
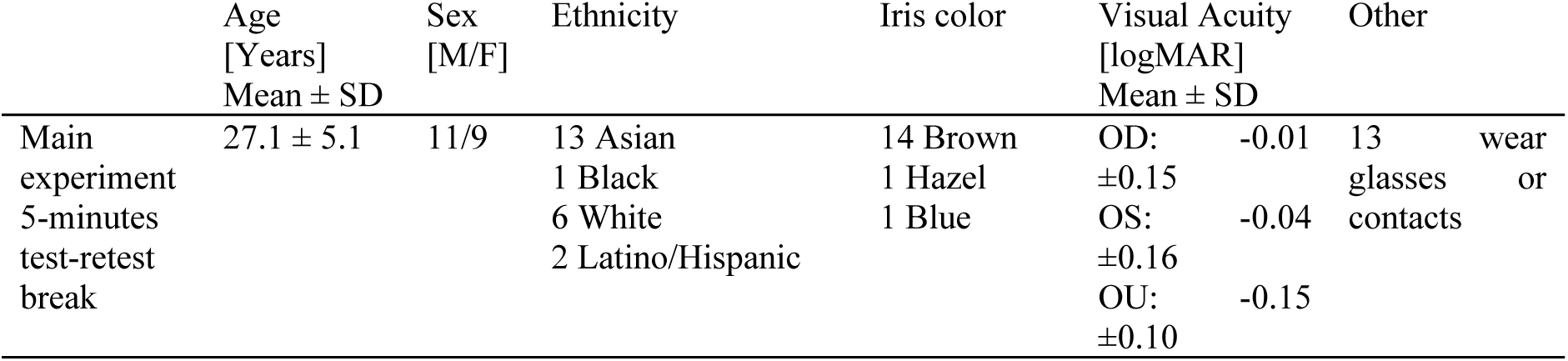
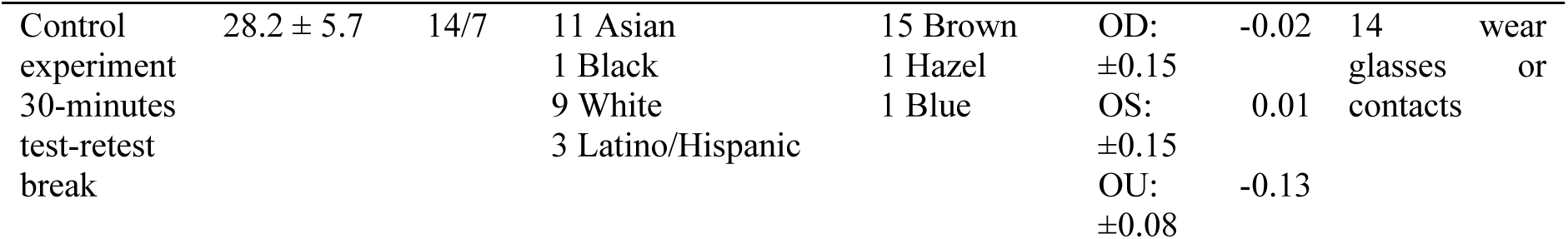
Demographic and ophthalmic screening summary for participants from both the 5-minute and the 30-minute experiments.

**Table 2:** Results for Swinging Flash-Light test in which an X indicates the presence of bias towards the retest and an O indicates the lack of bias towards the retest. Each parameter for the SFL setting is shown with the right eye and left eyes separated down the column and consensual and direct also separated. The first results show the 5-minute waiting period while the second showed the 30-minute rest period. Detailed reports of outcomes including numerical coefficients of repeatability, biases and basic statistics, are shown in Appendix Table 1.

| Parameter | Test-retest interval | OD Direct | OD Consensual | OS Direct | OS Consensual |
| --- | --- | --- | --- | --- | --- |
| Amplitude of Constriction [mm] | 5 min | o | X | X | o |
|  | 30 min | o | o | o | o |
| Latency of Onset of Constriction [msec] | 5 min | o | X | X | o |
|  | 30 min | o | o | o | X |
| Latency of Maximum Constriction [msec] | 5 min | X | o | X | X |
|  | 30 min | o | X | o | X |
| Velocity of Constriction [mm/sec] | 5 min | o | X | X | o |
|  | 30 min | o | o | o | o |
| Velocity of Relaxation [mm/sec] | 5 min | X | o | X | X |
|  | 30 min | o | o | o | o |

**Table 3:** Table showing results for Low-High Luminance test in which an X indicates the presence of bias towards the retest and an O indicates the lack of bias towards the retest. Each parameter for the Low-High Luminance setting is shown with the right eye and left eyes separated down the column. The first results show the 5-minute waiting period while the second showed the 30-minute rest period. Detailed reports of outcomes including numerical coefficients of repeatability, biases and basic statistics, are shown in Appendix Table 2.

| Parameter | Test-retest interval | OD | OS |
| --- | --- | --- | --- |
| Mean ($\mu$) pupillary diameter under Low Luminance [mm] | 5 min | O | O |
|  | 30 min | O | O |
| Mean ($\mu$) pupillary diameter under High Luminance [mm] | 5 min | X | X |
|  | 30 min | O | O |
| Relative Change in pupillary diameter [%] | 5 min | X | X |
|  | 30 min | O | O |
| Standard Deviation ($\sigma$) of pupillary diameter under Low Luminance [mm] | 5 min | X | X |
|  | 30 min | O | X |
| Standard Deviation ($\sigma$) of pupillary diameter under High Luminance [mm] | 5 min | O | O |
|  | 30 min | O | O |
| Low-High Difference in pupillary diameter [mm] | 5 min | X | X |
|  | 30 min | O | O |

To illustrate the analysis, example plots for the SFL experiment (Figure 1), and the Low-High Luminance experiment (Figure 2) are also shown.

The significant mean bias toward the retest for the SFL assay after a 5-minute break duration was reduced for the cohort that had a 30-minute break. On the other hand, the Coefficient of Repeatability (CoR) and mean bias values tended to be more variant and further away from zero, respectively (see Appendix Table 1). The absolute mean biases were compared between the 30- and 5-minute conditions. Only 2/21 or 9.5% of CoRs were more negative after 30 minutes, indicating more variance after a 30-minute break. Interestingly, 6/21 (28.5%) absolute mean biases were closer to zero after a 30-minute break. Since the 30-minute cohort included several new participants (n=6) and participants (n=15) that performed both tasks, we re-ran the analysis but only with the participants that took part in both experimental sessions to investigate whether the variance was affected by the cohort difference. We found that 11/21(52%) CoR values that were closer after a 30-minute break and 9/21 (43%) mean biases that were closer to zero after a 30-minute break.

The Low-High Luminance test’s results showed no significant mean bias, except in one setting of standard deviation for the low, after a 30-minute break. When comparing the 30-minute results with results after a 5-minute break, 66.7% and 58.3% of absolute mean biases and CoRs, respectively, also improved (Appendix Table 2), i.e., the mean biases were closer to zero and the CoR values were more negative, indicating less repeatability-related variability of the data. Additionally, we performed a linear correlation analysis for the first run’ data to test whether interocular visual acuity difference affected the RAPD score, but found no significant association (t= -0.13, p > 0.05). The Low-High Luminance test provides a low-to-high luminance difference measure, which we also correlated with interocular visual acuity difference. Visual acuity did also not correlate significantly with visual acuity of either left (t= 0.00, p>0.05) or right eye (t= 0.21, p>0.05).

## Discussion

Automated pupillography enables experimenter independent assessment of pupil light reflex kinetics, thereby generating computerized performance models for every trial. The current study’s main goal was to assess the repeatability of a clinical pupillography device, specifically its two integrated methods namely the SFL and Low-High Luminance tests. The results of the SFL test showed an overall trend towards a retest bias in most parameters with just a 5-minute test-retest interval (Table 2, Appendix Table 1). Zheng et al. validated a previous version of the device^23^ using intraclass correlations (ICCs) to assess repeatability and found that the outcome parameters showed ICCs ranging between 0.65 to 0.90. Although they did not use Bland-Altman analyses, they reported that the means of each parameter were systematically reduced across three trials, which suggests a re-test bias. Zheng et al. did not standardize the duration between test and retest, reporting that the interval varied by more than 5 minutes and elsewhere reporting that the interval ranged from 10 to 20 minutes.^23^ The retest bias after 5-minutes for the SFL test found in our study and the trend reported in Zheng et al.’s study may have been due to a lack of recovery time. Hence, we performed a control experiment with 30-minutes to test whether the retest biases would disappear and found that most biases were indeed eliminated for the SFL as well as the Low-High Luminance assays when the test retest interval was 30 minutes Interestingly the CoR values for each parameter during the SFL increased, indicating more variability after 30-minutes. As the 30-minute experiment included participants that took part in the 5-minute protocol as well, we re-ran the analysis of this subset of participants to check whether the variability was due to different groups. We found for the Low-High Luminance task that 67% (8/12) of the entire control cohort had smaller CoR values and 83% (10/12) for the subgroup that took part in both experiments (Appendix Tables 2 and 3). For the Swinging-Flash-Light method, the control experiment showed only 29% (6/21) of CoR values were smaller after the 30-minute break and when re-analyzing the subgroup of 15 participants that took part in both experiments only 14% (3/21) had smaller CoR (Appendix Tables 1 and 3). We conclude that for the Low-High Luminance test, 30-minute breaks eliminated retest bias and also reduced test-retest variability. For the SFL method on the other hand, retest bias was eliminated after 30-minute breaks however greater test-retest variability was introduced. Repeatability results of another study for a different device showed also a wide range of retest agreement i.e., ICCs from -0.2 to 0.8.^24^

Notably, the retest bias was most pronounced in the Low-High Luminance test, indicating that specific test settings may be more susceptible to temporal influences, likely due to the 5 seconds of continuous, bright white light exposure that caused prolonged photostress and thus delayed recovery compared to the SFL protocol and its eight 167millisecond flashes that were separated by 2.1 seconds of recovery phases. These results underscore the importance of considering the timing of repeat measurements when utilizing the EyeKinetix in clinical or research settings. To ensure reliable and reproducible outcomes, a minimum interval of at least 30-minutes is indicated between tests for both Eyekinetix methods. The reduction of retest bias as a function of test-retest break duration suggests that photostress induced during each trial and the recovery thereafter to baseline was the driver of the observer the retest bias. Moreover, although the retest bias per parameter was also reduced for the Low-High Luminance method, some mean bias still persisted. Given its 5 second white light exposure, it is plausible that of the two methods, the Low-High Luminance test is more affected by photostress-recovery than the SFL method.

It is noteworthy to list the current study’s limitations. While breaks between tests and retests were constant, time of day, and ambient noise were not. These two factors have been shown to affect the results of a pupillometry device^28,29^, potentially inducing confounding factors into the data. On the other hand, these factors are part of the clinical environment for which the devices are purposefully built. The current study tested healthy, young participants. Future studies may test the repeatability of older cohorts or cohorts with ophthalmic conditions. Some participants wore contact lenses with their correction whereas other participants performed experiments without best correction. A linear correlation was performed, specifically correlating interocular outcome measures of both tests with interocular visual acuity difference. The results showed no significant effect of visual acuity on the pupillary outcomes for both tests. One study of a pupillometer reported that the device reduced retest bias and less variability compared standard manual assessment of a trained clinician.^27^ A future study may repeat that protocol using the Eyekinetix by comparing its repeatability with manually gathered results to investigate whether the Eyekinetix generates also less retest bias and variability. Whether 30-minutes are sufficient for older patients or clinically affected cohorts is unclear and hence invites future research. Pupillography is a relatively new field with many different uses in the ophthalmology clinic besides the detection of RAPDs.^10^ However, clinicians often are under a time constraint with patients and a 30-minute waiting period where the patient cannot be dilated is not conducive to clinical flow. It is then recommended that staff members be appropriately trained in the device so that the need for retesting is minimized, reducing the clinical burden of this confounding factor. A future study may repeat that protocol using the Eyekinetix by comparing its repeatability with manually gathered results to investigate whether the Eyekinetix generates also less retest bias and variability.

## Conclusions

In this study, we assessed the repeatability of a clinical pupillography system in young, healthy participants. The primary aim was to evaluate the test-retest repeatability of the device in measuring the relative afferent pupillary defect (RAPD) by using a SFL approach and the Low-High Luminance variation in pupillary diameter for test-retest intervals of 5and 30-minutes. The key findings demonstrated retest bias after a 5-minute interval for SFL and Low-High luminance tests. **However, RAPD assessment, which is arguably the most useful metric provided by this form of pupillography, introduced bias when test-retest intervals were 5 or 30 minutes.** The retest bias was resolved when measurements were repeated after 30-minutes for the SFL method and significantly reduced for the Low-High Luminance method. After a 30-minute break, the SFL setting showed more variability whereas the Low-High Luminance test results showed less variability. These results suggest that pupillography assay is repeatable with a 30-minute break between test and retest.

## Data Availability

All relevant data are within the manuscript and its Supporting Information files.

## Acknowledgments

The authors acknowledge that Konan Medical provided the EyeKinetix device at no cost to the UC Irvine Department of Ophthalmology in exchange for the predecessor instrument, RAPDx, which was also manufactured and distributed by Konan Medical and previously purchased by the UC Irvine Department of Ophthalmology.

## Authors contributions

**Conceptualization**: All authors

**Data Curation**: Jacob Diaz

**Formal Analysis**: Jacob Diaz, Jan Skerswetat

**Funding Acquisition**: Andrew Browne

**Investigation**: Jacob Diaz

**Methodology**: All authors

**Project Administration:** Andrew Browne

**Resources:** Andrew Browne

**Software:** Jan Skerswetat

**Supervision:** Andrew Browne

**Validation:** All authors

**Visualization:** Jacob Diaz, Jan Skerswetat

**Writing – Original Draft Preparation:** Jacob Diaz, Jan Skerswetat

**Writing – Review and Editing:** All authors

**Funding:** Andrew Browne

## Supplemental Tables

**Supplemental Table 1:** Summary of basic and Bland-Altman statistics for the Swinging-Flash-Light approach. The results for the 30-minute break interval condition are in italic font below each 5-minute results. Means (µ), standard deviations (σ), Coefficients of Repeatability (CoR), Mean and Proportional Biases (Prop. Bias) are shown. Parameters are: Relative Afferent Pupillary Defect (RAPD) score, Amplitude of Constriction (AC), Latency of Onset-Constriction (LoC), Latency of Peak Constriction (LPC), Velocity of Constriction (VC), and Velocity of Recovery (VR). *\*highlights p-values which are smaller than 0.05; ̈ indicates CoR results for 30-minutes that are more negative than results during the 5-minute break interval; ̈̈ indicates absolute mean bias results for the 30-minute break interval that are smaller than results during the 5-minute break interval*.

| SFL Parameters Results | Basic Statistics |  | Bland-Altman Statistics |  |  |  |
| --- | --- | --- | --- | --- | --- | --- |
| | Test<br>$\mu \pm \sigma$ | Retest<br>$\mu \pm \sigma$ | CoR | Mean Bias | Mean Bias p | Prop. Bias p |
| **RAPD Score** (5min break) | -0.09 ± 0.23 | -0.16 ± 0.31 | ± 0.85 | 0.07 | p= 0.30 | p= 0.18 |
| 30min break | -0.18 ± 0.28 | -0.16 ± 0.32 | ± 0.59 | -0.01 | p= 0.81 | p= 0.29 |
| **AC OD Direct [mm]** | 1.33 ± 0.22 | 1.34 ± 0.22 | ± 0.25 | -0.008 | p= 0.69 | p= 0.72 |
| | 1.29 ± 0.22 | 1.33 ± 0.18 | ± 0.38 | -0.04 | p= 0.19 | p= 0.38 |
| **AC OD Consensual [mm]** | 1.31 ± 0.21 | 1.34 ± 0.22 | ± 0.16 | -0.03 | p= 0.03* | p= 0.42 |
| | 1.30 ± 0.22 | 1.30 ± 0.21 | ± 0.39 | -0.02 | p= 0.44 | p= 0.48 |
| **AC OS Direct [mm]** | 1.36 ± 0.22 | 1.39 ± 0.24 | ± 0.15 | -0.04 | p= 0.008** | p= 0.14 |
| | 1.35 ± 0.20 | 1.38 ± 0.17 | ± 0.40 | -0.04 | p= 0.22 | p= 0.63 |
| **AC OS Consensual [mm]** | 1.28 ± 0.22 | 1.29 ± 0.24 | ± 0.24 | -0.01 | p= 0.51 | p= 0.45 |
| | $1.24 \pm 0.22$ | $1.26 \pm 0.17$ | $\pm 0.37$ | -0.03 | $p = 0.33$ | $p = 0.37$ |
| **LoC OD Direct [msec]** | $239.0 \pm 14.8$ | $234.7 \pm 11.1$ | $\pm 25.9$ | 4.25 | $p = 0.06$ | $p = 0.07$ |
| | $240.6 \pm 16.6$ | $239.1 \pm 19.6$ | $\pm 36.5$ | 5.48 | $p = 0.07$ | $p = 0.99$ |
| **LoC OD Consensual [msec]** | $238.1 \pm 16.5$ | $232.2 \pm 10.7$ | $\pm 34.4$ | 5.90 | $p = 0.05$ | $p = 0.02^*$ |
| | $240.7 \pm 18.1$ | $237.4 \pm 19.3$ | $\pm 29.1$ | 3.95 | $p = 0.10$ | $p = 0.58$ |
| **LoC OS Direct [msec]** | $233.1 \pm 11.3$ | $228.9 \pm 10.1$ | $\pm 20.5$ | 4.15 | $p = 0.02^*$ | $p = 0.43$ |
| | $237.3 \pm 15.8$ | $237.4 \pm 17.6$ | $\pm 25.2$ | 1.57 | $p = 0.44$ | $p = 0.78$ |
| **LoC OS Consensual [msec]** | $238.2 \pm 14.6$ | $237.2 \pm 12.3$ | $\pm 23.5$ | 0.95 | $p = 0.62$ | $p = 0.23$ |
| | $247.3 \pm 18.3$ | $239.0 \pm 16.1$ | $\pm 25.6$ | 4.76 | $p = 0.03^*$ | $p = 0.06$ |
| **LC OD Direct [msec]** | $712.2 \pm 51.4$ | $698.0 \pm 53.5$ | $\pm 50.3$ | 14.15 | $p = 0.003^{**}$ | $p = 0.61$ |
| | $725.4 \pm 29.2$ | $722.1 \pm 52.2$ | $\pm 73.2$ | 11.10 | $p = 0.07$ | $p = 0.20$ |
| **LC OD Consensual [msec]** | $698.1 \pm 48.6$ | $692.2 \pm 44.4$ | $\pm 50.2$ | 5.85 | $p = 0.17$ | $p = 0.32$ |
| | $717.1 \pm 24.4$ | $705.6 \pm 34.3$ | $\pm 56.8$ | 14.14 | $p = 0.005^{**}$ | $p = 0.76$ |
| **LC OS Direct [msec]** | $710.5 \pm 50.8$ | $702.1 \pm 46.7$ | $\pm 45.9$ | 8.35 | $p = 0.04^*$ | $p = 0.28$ |
| | $732.1 \pm 27.7$ | $728.7 \pm 27.6$ | $\pm 52.2$ | 3.29 | $p = 0.43$ | $p = 0.25$ |
| **LC OS Consensual [msec]** | $695.6 \pm 51.1$ | $684.7 \pm 48.2$ | $\pm 52.5$ | 10.90 | $p = 0.02^*$ | $p = 0.51$ |
| | $713.6 \pm 35.8$ | $704.0 \pm 40.0$ | $\pm 59.3$ | 13.33 | $p = 0.01^*$ | $p = 0.43$ |
| **VC OD Direct [mm/sec]** | $4.98 \pm 0.73$ | $5.06 \pm 0.67$ | $\pm 0.73$ | -0.07 | $p = 0.22$ | $p = 0.23$ |
| | $4.73 \pm 0.80$ | $4.93 \pm 0.81$ | $\pm 1.57$ | -0.24 | $p = 0.07$ | $p = 0.89$ |
| **VC OD Consensual [mm/sec]** | $4.93 \pm 0.66$ | $5.10 \pm 0.71$ | $\pm 0.54$ | -0.18 | $p < 0.001^{***}$ | $p = 0.31$ |
| | $4.81 \pm 0.84$ | $4.90 \pm 0.92$ | $\pm 1.44$ | -0.21 | $p = 0.08$ | $p = 0.85$ |
| **VC OS Direct [mm/sec]** | $5.00 \pm 0.65$ | $5.22 \pm 0.74$ | $\pm 0.61$ | -0.22 | $p < 0.001^{***}$ | $p = 0.09$ |
| | $4.90 \pm 0.82$ | $4.98 \pm 0.71$ | $\pm 1.47$ | -0.23 | $p = 0.06$ | $p = 0.83$ |
| **VC OS Consensual [mm/sec]** | $4.87 \pm 0.74$ | $5.00 \pm 0.75$ | $\pm 0.79$ | -0.11 | $p = 0.10$ | $p = 0.90$ |
| | $4.65 \pm 0.84$ | $4.77 \pm 0.77$ | $\pm 1.47$ | -0.19 | $p = 0.11$ | $p = 0.60$ |
| VR OD Direct [mm/sec] | 1.75 ± 0.30 | 1.78 ± 0.25 | ± 0.33 | -0.04 | p= 0.16 | p= 0.04* |
|  | *1.71 ± 0.33* | *1.71 ± 0.34* | *± 0.54* | *-0.05* | *p= 0.27* | *p= 0.92* |
| VR OD Consensual [mm/sec] | 1.76 ± 0.30 | 1.80 ± 0.27 | ± 0.32 | -0.04 – | p= 0.12 | p= 0.24 |
|  | *1.73 ± 0.30* | *1.75 ± 0.33* | *± 0.58* | *0.07* | *p= 0.13* | *p= 0.47* |
| VR OS Direct [mm/sec] | 1.78 ± 0.26 | 1.82 ± 0.28 | ± 0.27 | -0.05 | p= 0.04* | p= 0.37 |
|  | *1.71 ± 0.29* | *1.73 ± 0.28* | *± 0.62* | *-0.06* | *p= 0.24* | *p= 0.43* |
| VR OS Consensual [mm/sec] | 1.75 ± 0.32 | 1.78 ± 0.25 | ± 0.38 | -0.03 | p= 0.41 | p= 0.02* |
|  | *1.70 ± 0.35* | *1.71 ± 0.30* | *± 0.53* | *-0.05* | *p= 0.26* | *p= 0.31* |
| “30min < 5min [n/%] |  |  | “2/21 | ““6/21 |  |  |
| ““30min closer to 0 than 5min [n/%] |  |  | “9.5% | ““28.5% |  |  |

**Supplemental Table 2:**
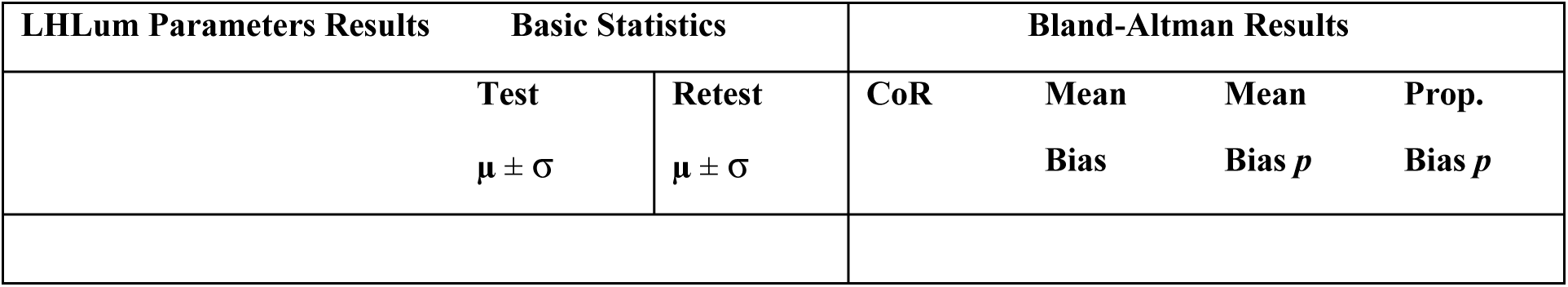
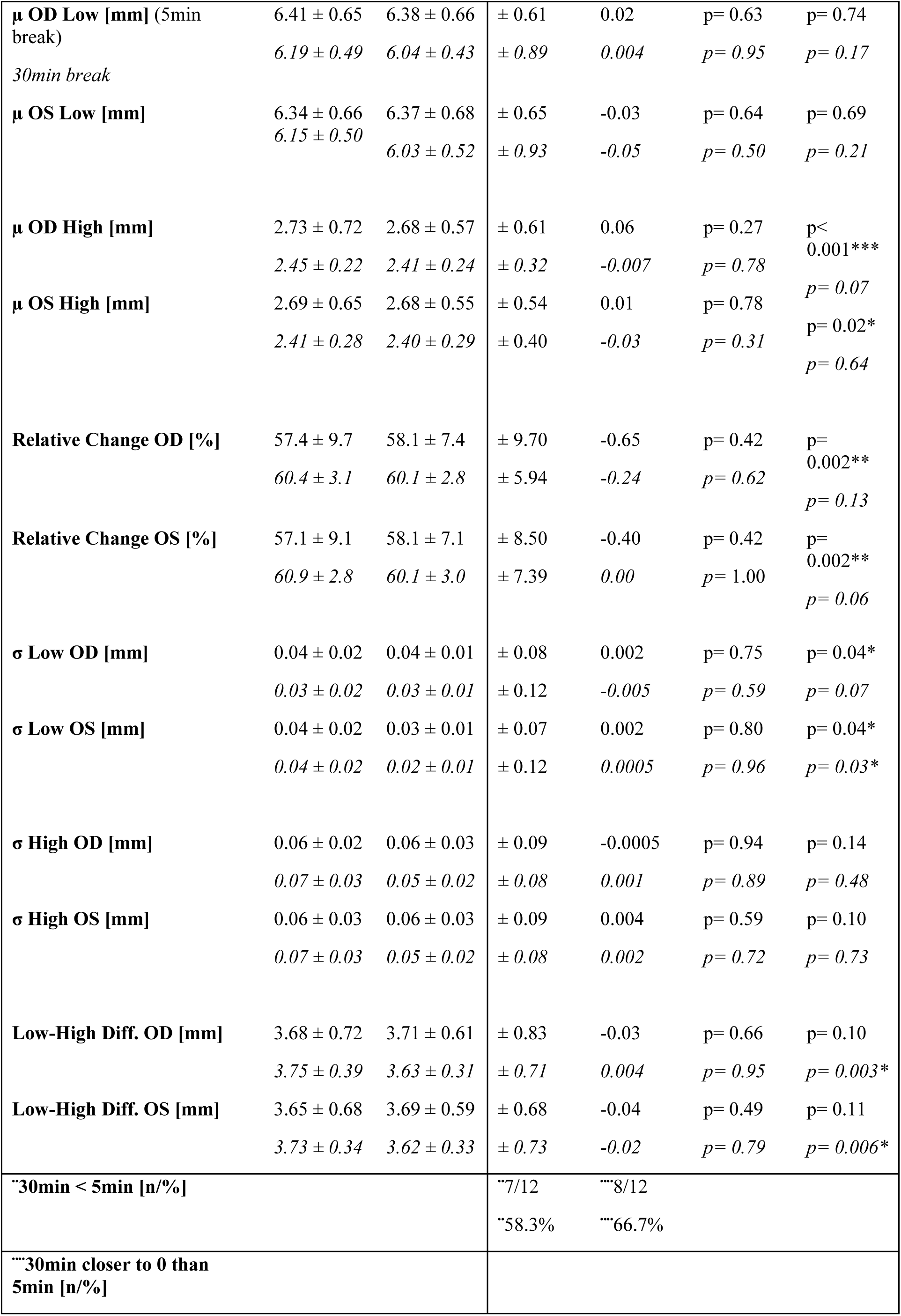
Low-High Luminance Test results summary for basic statistics mean (m) and standard deviations (σ) and Bland-Altman statistics. The results for the 30-minute break interval condition are in italic font below each 5-minute result. Means (m), standard deviations (σ), Coefficients of Repeatability (CoR), Mean and Proportional Biases (Prop. Bias) are shown. Parameters are: Mean (µ) Low, Mean (µ) High, Relative Change, Standard Deviation (σ) High, Standard Deviation (σ) Low, and Low-High Difference. *\* highlights p-values which are smaller than 0.05; ̈ indicates CoR results for 30-minutes that are more negative than results during the 5 minute break interval; ̈̈ indicates absolute mean bias results for the 30-minute break interval that are smaller than results during the 5-minute break interval*.

**Supplemental Table 3:** Control analysis for both SFL and LHLum test for 15 participants that took part in both experiments.

| SFL Parameters Results | Basic Statistics |  | Bland-Altman Statistics |  |  |  |
| --- | --- | --- | --- | --- | --- | --- |
| | Test $\mu \pm \sigma$ | Retest $\mu \pm \sigma$ | CoR | Mean Bias | Mean Bias p | Prop. Bias p |
| RAPD Score (5min)<br>(30min) | -0.12 ± 0.24 | -0.15 ± 0.33 | 0.63 | 0.04 | 0.21 | 0.65 |
| | -0.11 ± 0.25 | -0.08 ± 0.30 | 0.48 | -0.03 | 0.46 | 0.68 |
| AC OD Direct [mm] | 1.29 ± 0.24 | 1.30 ± 0.22 | 0.20 | -0.01 | 0.51 | 0.83 |
| | 1.26 ± 0.23 | 1.31 ± 0.18 | 0.26 | -0.05 | 0.11 | 0.13 |
| AC OD Consensual [mm] | 1.28 ± 0.23 | 1.30 ± 0.24 | 0.09 | -0.02 | 0.72 | 0.15 |
| | 1.25 ± 0.23 | 1.28 ± 0.20 | 0.26 | -0.03 | 0.33 | 0.41 |
| AC OS Direct [mm] | 1.32 ± 0.22 | 1.34 ± 0.23 | 0.08 | -0.02 | 0.47 | 0.08 |
| | 1.29 ± 0.24 | 1.33 ± 0.19 | 0.24 | -0.04 | 0.11 | 0.19 |
| AC OS Consensual [mm] | 1.24 ± 0.22 | 1.25 ± 0.22 | 0.19 | -0.01 | 1.00 | 0.83 |
| | 1.21 ± 0.24 | 1.25 ± 0.17 | 0.24 | -0.04 | 0.02* | 0.22 |
| LoC OD Direct [msec] | 238.93 ± 15.26 | 235.53 ± 11.24 | 18.68 | 3.40 | 0.10 | 0.19 |
| | 240.00 ± 14.05 | 235.73 ± 18.01 | 22.87 | 4.27 | 0.19 | 0.18 |
| LoC OD Consensual [msec] | 236.73 ± 18.39 | 233.33 ± 11.64 | 25.27 | 3.40 | 0.03* | 0.32 |
| | 239.00 ± 16.45 | 234.60 ± 17.64 | 19.44 | 4.40 | 0.66 | 0.11 |
| **LoC OS Direct [msec]** | 232.27<br>± 12.48<br>236.73<br>± 14.72 | 230.00<br>± 10.49<br>235.67<br>± 15.64 | 11.72<br>14.97 | 2.27<br>1.07 | 0.21<br>0.66 | 0.16<br>0.60 |
| **LoC OS Consensual [msec]** | 235.67<br>± 15.64<br>243.40<br>± 17.41 | 237.80<br>± 13.65<br>236.73<br>± 14.72 | 11.03<br>16.58 | -2.13<br>6.67 | 0.19<br>0.23 | 0.16<br>0.01* |
| **LC OD Direct [msec]** | 703.80<br>± 46.47<br>722.67<br>± 39.56 | 688.33<br>± 51.39<br>711.60<br>± 52.94 | 36.09<br>54.58 | 15.47<br>11.07 | 0.33<br>0.06 | 0.01*<br>0.15 |
| **LC OD Consensual [msec]** | 690.60<br>± 40.95<br>711.53<br>± 35.36 | 682.87<br>± 40.64<br>699.53<br>± 40.66 | 34.59<br>39.74 | 7.73<br>12.00 | 0.95<br>0.33 | 0.11<br>0.04* |
| **LC OS Direct [msec]** | 702.73<br>± 46.69<br>723.87<br>± 41.03 | 694.93<br>± 46.48<br>718.20<br>± 42.78 | 32.29<br>27.21 | 7.80<br>5.67 | 0.96<br>0.65 | 0.09<br>0.14 |
| **LC OS Consensual [msec]** | 686.20<br>± 45.09<br>712.60<br>± 45.49 | 677.20<br>± 47.52<br>699.53<br>± 44.24 | 32.38<br>37.28 | 9.00<br>13.07 | 0.60<br>0.81 | 0.05<br>0.02* |
| **VC OD Direct [mm/sec]** | 4.89 ±<br>0.75<br>4.60 ±<br>0.81 | 4.96 ±<br>0.65<br>4.89 ±<br>0.71 | 0.53<br>1.17 | -0.06<br>-0.28 | 0.17<br>0.52 | 0.37<br>0.09 |
| **VC OD Consensual [mm/sec]** | 4.86 ±<br>0.72<br>4.63 ±<br>0.83 | 5.03 ±<br>0.75<br>4.84 ±<br>0.80 | 0.35<br>1.04 | -0.17<br>-0.21 | 0.50<br>0.87 | 0.00*<br>0.15 |
| **VC OS Direct [mm/sec]** | 4.92 ±<br>0.63<br>4.66 ±<br>0.85 | 5.11 ±<br>0.71<br>4.88 ±<br>0.66 | 0.43<br>0.93 | -0.19<br>-0.22 | 0.17<br>0.12 | 0.00*<br>0.10 |
| **VC OS Consensual [mm/sec]** | 4.82 ±<br>0.70<br>4.50 ±<br>0.85 | 4.87 ±<br>0.68<br>4.72 ±<br>0.68 | 0.56<br>1.00 | -0.05<br>-0.22 | 0.73<br>0.19 | 0.49<br>0.12 |
| **VR OD Direct [mm/sec]** | 1.71 ±<br>0.29 | 1.75 ±<br>0.24 | 0.22 | -0.05 | 0.10 | 0.14 |
|  | 1.67 ±<br>0.29 | 1.72 ±<br>0.29 | 0.41 | -0.05 | 0.97 | 0.36 |
| **VR OD Consensual**<br>**[mm/sec]** | 1.73 ±<br>0.31<br>1.69 ±<br>0.27 | 1.77 ±<br>0.28<br>1.74 ±<br>0.28 | 0.21<br>0.28 | -0.04<br>-0.05 | 0.30<br>0.73 | 0.17<br>0.20 |
| **VR OS Direct [mm/sec]** | 1.74 ±<br>0.25<br>1.68 ±<br>0.27 | 1.77 ±<br>0.27<br>1.71 ±<br>0.24 | 0.18<br>0.28 | -0.04<br>-0.03 | 0.42<br>0.61 | 0.15<br>0.38 |
| **VR OS Consensual**<br>**[mm/sec]** | 1.73 ±<br>0.30<br>1.66 ±<br>0.31 | 1.74 ±<br>0.23<br>1.71 ±<br>0.26 | 0.25<br>0.37 | -0.01<br>-0.05 | 0.03*<br>0.30 | 0.71<br>0.30 |
| **LHLum Parameters**<br>**Results** | **Basic**<br>**Statistic**<br>**s** |  | **Bland-**<br>**Altman**<br>**Statistic**<br>**s** |  |  |  |
|  | **Test**<br>**μ ± σ** | **Retest**<br>**μ ± σ** | **CoR** | **Mean**<br>**Bias** | **Mean**<br>**Bias p** | **Prop.**<br>**Bias p** |
| **μ OD Low [mm]** | 6.30 ±<br>0.67<br>6.23 ±<br>0.68 | 6.25 ±<br>0.66<br>6.16 ±<br>0.67 | 0.48<br>0.48 | 0.05<br>0.07 | 0.90<br>0.96 | 0.46<br>0.26 |
| **μ OS Low [mm]** | 6.25 ±<br>0.68<br>6.18 ±<br>0.64 | 6.28 ±<br>0.68<br>6.12 ±<br>0.66 | 0.50<br>0.51 | -0.03<br>0.06 | 0.99<br>0.78 | 0.64<br>0.41 |
| **μ OD High [mm]** | 2.71 ±<br>0.83<br>2.50 ±<br>0.31 | 2.64 ±<br>0.65<br>2.50 ±<br>0.37 | 0.48<br>0.25 | 0.07<br>0.00 | 0.00 *<br>0.07 | 0.27<br>0.94 |
| **μ OS High [mm]** | 2.67 ±<br>0.74<br>2.46 ±<br>0.33 | 2.66 ±<br>0.62<br>2.47 ±<br>0.37 | 0.42<br>0.27 | 0.01<br>-0.01 | 0.02 *<br>0.28 | 0.91<br>0.84 |
| **Relative Change OD [%]** | 57.00 ±<br>11.27<br>59.80 ±<br>3.53 | 57.80 ±<br>8.56<br>59.33 ±<br>3.50 | 7.74<br>2.21 | -0.80<br>0.47 | 0.00 *<br>0.92 | 0.45<br>0.13 |
| **Relative Change OS [%]** | 57.40 ±<br>10.54<br>60.13 ±<br>3.54 | 57.80 ±<br>8.20<br>59.60 ±<br>3.48 | 6.62<br>2.76 | -0.40<br>0.53 | 0.00 *<br>0.87 | 0.65<br>0.16 |
| **σ Low OD [mm]** | 0.04 ±<br>0.02<br>0.03 ±<br>0.02 | 0.03 ±<br>0.01<br>0.03 ±<br>0.01 | 0.05<br>0.04 | 0.01<br>0.00 | 0.03 *<br>0.29 | 0.11<br>0.91 |
| **σ Low OS [mm]** | 0.04 ±<br>0.02<br>0.03 ±<br>0.02 | 0.03 ±<br>0.01<br>0.02 ±<br>0.01 | 0.05<br>0.03 | 0.01<br>0.01 | 0.09<br>0.34 | 0.26<br>0.05 |
| **σ High OD [mm]** | 0.05 ±<br>0.03<br>0.06 ±<br>0.03 | 0.06 ±<br>0.04<br>0.05 ±<br>0.03 | 0.06<br>0.05 | -0.01<br>0.01 | 0.19<br>0.58 | 0.54<br>0.13 |
| **σ High OS [mm]** | 0.06 ±<br>0.02<br>0.06 ±<br>0.03 | 0.06 ±<br>0.04<br>0.05 ±<br>0.03 | 0.07<br>0.04 | 0.00<br>0.01 | 0.14<br>0.73 | 0.78<br>0.73 |
| **Low-High Diff. OD [mm]** | 3.59 ±<br>0.81<br>3.73 ±<br>0.50 | 3.61 ±<br>0.65<br>3.65 ±<br>0.44 | 0.66<br>0.31 | -0.02<br>0.08 | 0.07<br>0.15 | 0.79<br>0.07 |
| **Low-High Diff. OS [mm]** | 3.58 ±<br>0.76<br>3.71 ±<br>0.46 | 3.62 ±<br>0.63<br>3.64 ±<br>0.43 | 0.53<br>0.33 | -0.04<br>0.07 | 0.07<br>0.58 | 0.58<br>0.15 |

